# Covid-19 High Attack Rate Can Lead to High Case Fatality Rate

**DOI:** 10.1101/2021.03.23.21254184

**Authors:** Tareef Fdahil Raham

## Abstract

**Background:** During the current Covid-19 pandemic case fatality rate (CFR) estimates were subjected to a lot of debates regarding the accuracy of its estimations, predictions, and the reason of across countries variances. In this context, we conduct this study to see the relationship between attack rate (AR) and CFR.

The study hypothesis is based on two: 1-evidence suggests that the mortality rate (MR) has a positive influence on case fatality ratio (CFR), 2- and increase number of Covid-19 cases leads to increased mortality rate (MR).

**Material and methods:** Thirty countries and territories were chosen. Inclusion criterion was > 500 Covid-19 reported cases per 10,000 population inhabitants. Data on covid-19 cases and deaths was selected as it was on March 10, 2021. Statistical methods used are descriptive and one-sample Kolmogorov-Smirnov (K-S), the one-way ANOVA, Levene, least significant different (LSD), and matched paired-samples T-tests.

**Results:** ANOVA test showed a significant difference at P<0.01 among all studied groups concerning AR and CFR mean values. Group of countries with MR ≥ 15 death / 10^4^ inhabitants recorded the highest level of crude mean CFR and AR values, and recorded the highest gap with leftover groups, especially with countries reported MR of <10 death/ 10^4^ inhabitants. There were independence 95% confidence intervals of mean CFR and AR values between countries with ≥ 15 death / 10^4^ MR and countries with MR of <10 death /10^4^. There was a significant difference between countries with MR ≥ 15 death / 10^4^ inhabitants and countries with MR of <10 death / 10 ^4^ inhabitants groups through least significant difference (LSD) test for CFR%(0.042 p-value) and Games Howell (GH) test for AR/10^4^ (p-value 0.000).

**Conclusions:** Total and mean AR and CFR are higher in high MR countries compared to low MR countries.

## Introduction

The case fatality rate (CFR) sometimes is called case fatality risk or case fatality ratio. The CFR is used to define the probability that a case dies from the infection.^1^

SARS-CoV-2 cluster infections were recognized as a critically important issue regarding preventing controlling its spreading. Examples of these clusters include family clusters (which is the dominant type)^2^, nosocomial infections^3^, transmission during gathering settings (like a birthday party, shopping malls, conferences, religious gatherings, offices, tourists, prisons, and funeral).^4^ Of the many driving factors of the strong transmissibility, cluster infections play critical roles in the widespread of disease and exponentially increases the number of cases .^2^ It is thought that super-spreaders of COVID-19 play a role in transmission within these clusters since it is partially contribute to the high transmission risk of SARS-CoV-2 . ^2^ Understanding cluster infections of SARS-CoV-2 transmission of the disease is an important strategy for prevention and control measures to contain the Covid-19 pandemic.^2^

High fatality rates were also reported within these SARS-CoV-2 cluster infections. ^3^

At country levels CFR was seen to be different in different places and seen to be not a constant finding furthermore, it can decrease or increase over time. ^5^ By 1st February, it was 5.8% and greater than 20% in the center of the outbreak in Wuhan while it was 0.7% across the rest of China.^1,2^ The rate reported outside China in February was even lower.^6^ The highest rates were found in West and North Europe (14%–19%), and North America (9%–12%).^6^

During a disease outbreak, estimation of the (CFR) is usually used as an indication of its severity, and is useful for planning and determining the intensity of a response to an outbreak as a guide to plan public health strategies.^1,7, 8^

Studies have reported multiple factors for variances in COVID-19 CFR among countries, these include demographics & social factors, comorbidities, and environmental factors (such as temperature, humidity, and air pollution).^9^ Although high attack rates (AR)s were studied in small clusters and small locations the underlying reason for such variances was not identified, while high mortalities during wave peaks were attributed to system failure to cope with increases burden . There is evidence that CFR is high among countries with high MR.^10,11^ There is no previous literature proving that a high number of confirmed cases could lead to high CFR as far as we know. Previous literatures usually attribute high CFR to low estimates due to low testing. Another cause of high CFR in certain places is usually attributed to unknown or yet not yet proved causes. The relation between case overload and CFR was not studied at the country level in detail before. From a global health perspective, there is evidence of a knowledge gap in this research field aspect.

In this paper, we look for the role of national Covide-19 case overload (AR) in determining CFR.

The study aims to look for the relation between AR and CFR in different countries. This study will help in the development of prevention and intervention measures to fight against this global public health crisis.

## Material and methods

Thirty countries and territories were chosen. Inclusion criterion was > 500 Covid-19 reported cases per 10,000 population inhabitants. Data on covid-19 cases and deaths was selected as it was on March 10,2021. Countries and territories were classified into three group groups: group I: countries with mortality rate ≥ 15 death /10^4^ population inhabitants; group II: ≥ 10-15death / 10^4^ population inhabitants; and group III <10 death /10^4^ population inhabitants. Supplementary attached file contains original data, computed data, and references for data sources.

### Definitions

The detected attack rate(AR) for a given country was calculated as the total number of reported cases divided by the estimated population of that country. Crude COVID-19 CFR was calculated as the total number of COVID-19 deaths divided by the number of total COVID-19 confirmed cases by march 10,2021 multiplied by 100.

### Methodology

The following statistical data analysis approaches were used under the application of the statistical package (SPSS) ver. (22.0):

### Descriptive data analysis

Mean value, standard deviation, standard error, (95%) confidence interval, and graphical presentation by using Bar Charts. Mean values and the two extremes values (min. and max.) were computed assuming that data followed normal distribution function.

### Inferential data analysis

These were used to accept or reject the statistical hypotheses, which included the following:

a. The One-Sample Kolmogorov-Smirnov (K-S) test. This is a goodness-of-fit test whether the observations could reasonably have come from the specified distribution.
b. The One-Way ANOVA procedure to test the hypothesis that several means are equal. In addition to that, we applied after rejecting the statistical hypotheses, least significant difference (LSD) test requiring equal variances was assumed, and Games Howell test not requiring equal variances was assumed.
c. Levene test: was used to test homogeneity of variances for equality of variances of two and several independent groups.
d. Matched paired-samples T-Test procedure was used to compare the means of two variables for a single group. It computes the differences between values of the two variables for each case and tests whether the average differs from zero.

## Results and Findings

Group I countries showed higher total attack rate and higher total CFR% than group III countries. Group II countries showed the lowest test coverage among three groups (table1) .

Table No. (2) represent a one-sample “Kolmogorov-Smirnov” test procedure comparing the observed cumulative distribution function for studied data with a specified theoretical distribution, which proposed normal shape (i.e. bell shape), for the studied markers.

The results showed that the test’s distribution was normal for the studied reading’s markers since no significant levels were accounted (P-value >0.05). This enabled us of applying conventional two methods of statistics: the descriptive methods of estimations (points and intervals), and the inferential statistics.

Table (3) represents a summary statistic, such as mean values, standard deviation, standard error, 95% confidence interval concerning mean parameter of the studied population, and the two extreme values (minimum and maximum) of studied values for (CFR %, and AR / 10^4^ population inhabitants) markers. Results showed that the CFR % marker recorded a high level of mean value concerning group I with a high gap in relation to leftover groups, especially to group III, which accounted for the lowest level among all markers. In addition to that, first and third groups recorded an independent or non-interferer of 95% confidence interval for mean values for each other. Furthermore, group III recorded independent (non-interferer) of 95% confidence interval for mean values in relation to other groups.

Regarding the (AR / 10^4^) marker, results showed that group I recorded a high level of mean value, and a high gap in relation to leftover groups, especially to group III, which accounted for the lowest level for preceding markers. In addition to that, first and third groups recorded independent or non-interferer of 95% confidence interval for mean values.

Concerning testing the compound statistical hypothesis, which says that studied group’s concerning (CFR%, and AR/104) readings are thrown from the same population, and that should be proved according to of testing equal variances are assumed, as well as equal mean values are assumed through “Levene and one-way ANOVA” tests respectively, and as illustrated in the table (4).

Concerning testing equal variances of CFR% marker, the Levene test showed that no significant differences are accounted at P>0.05 among studied groups and a highly significant result (0.006 p-value) concerning AR. ANOVA test showed a significant difference at P<0.01 among all studied groups concerning AR and CFR mean values.

The alternative statistical hypothesis says that at least two groups are not equal due to their mean values. This was tested through the LSD test for CFR% marker, and Games Howell (GH) test for AR/104 marker (table5).

Results in table 5 showed that no significant difference between groups I, and II regarding both studied markers, and no significant difference between groups II and III regarding the AR marker. There were significant differences among the leftover comparisons in at least at P<0.05 for each of the studied markers. There was a significant difference between the I and III groups in both two tests.

**Table 5:** Multiple Comparisons using (LSD) and (GH) tests for studied markers among studied groups of countries regarding mortality rates

| Marker | (I) Group | (J) Group | Mean Diff. (I-J) | Sig. | C.S. (*) |
| --- | --- | --- | --- | --- | --- |
| CFR% | I | II | 0.391 | 0.066 | NS |
|  |  | III | 1.399 | 0.000 | HS |
|  | II | III | 1.008 | 0.000 | HS |
| AR/10<sup>4</sup> | I | II | 147.6 | 0.390 | NS |
|  |  | III | 244.0 | 0.042 | S |
|  | II | III | 96.4 | 0.424 | NS |

(\*) HS: Highly Sig. at P<0.01; S: Sig. at P<0.05; Non Sig. at P>0.05; Testing based on GH test.

## Discussion

Commentators may consider CFR as if it’s a steady and unchanging number confined to a specific disease in general and Covid-19 in particular. Although CFR may be a useful measure to assess the magnitude of a disease outbreak, it has been consistently being subjected to underestimation and overestimation. ^12^

Overestimate of CFR is largely due to underestimation ^12^of cases especially encountered in an infection with a range of manifestations from relatively mild to severe . 1

Several countries have implemented a strategy of massive screening, aiming at identifying positives^9^.Testing capacity, which is associated with the availability of resources and manpower, is the single most important factor that can tremendously affect the CFR. ^12^The lack of availability of widespread testing leads to an ascertainment bias toward severe cases.^13^ A more reliable method of estimating the magnitude of the outbreak would be an assessment of the infection fatality rate (IFR).^13,14^

Underestimation of death accounts can lead to erroneously low CFR. This could be due to: (1) in crude estimated, some patients encountered as non-dead are still hosted in intensive care units^9^, (2) deaths caused by COVID-19 may be misattributed to other death classifications and codes,^15^ and (3)deaths Confirmed counts are subject to time lags. ^3, 16^This means that reported with COVID-19 will die at a later date.^1, 3,14^

Our significant findings of a highly significant association between AR and CFR is supported by the following previous observations: (1) available data about South Americans and Asian countries that took the strictest measures, and they also had relatively lower COVID-19 CFR^6,17^, (2) it has been noticed that in European countries that have had both large numbers of cases and deaths^15^, the average of country-specific CFR was at 0.7% –1.3% at early times of pandemic raised to about 2%–3.31%□ thereafter^6^, this increase in Covid-19 CFR possibly goes in parallel with an increase in number of cases, (4) the CFR of COVID-19 differs by location, and has changed during the period of the outbreak ^3,6^, (5) small clusters of fatal COVID-19 infections were reported previously with high CFR within these clusters (families, tourists, long□term care hospitals and facilities, etc.).^18,19,20,21,22^ There were also identified “vulnerable” clusters of counties in USA with high mortality incidence ratio.^23^ In UK A few areas saw COVID-19 mortality more than seven times the expected level compared with the rest of the country^24^, (6)association between population size and COVID-19 CFR^6^, and (7) our findings were in concordance with recent studies which found very high positive significant correlation between total deaths/1M and the total number of cases and a very high positive influence of the COVID-19 MR on the CFR^10,11^.

We suggested that AR plays an important role in explaining variances.

The underlying cause for increased CFR with increased AR is possibly related to high viral overload as it was observed in clustering infections a phenomenon already described before that is characterized by mortality and fatality rates.

It was thought that CFR is used as a measure of disease severity and is often used for predicting disease course or outcome.

In our study, CFR reflects the density of infection and AR value as well as the severity of the disease. As far as the epidemiology concerns with the virulence of the disease in addition to the transmissibility of infectious disease, ^25^ our finding of the significant role of AR will add an important factor explaining various virulence of epidemics in different places and times.

### Conclusions

During Covid-19 pandemic, the CFR is a poor measure of the mortality risk of the disease. CFR is a reflection and consequence of AR, concepts regarding the severity of the disease should be directed to the ability to have high AR rather than to its high CFR since CFR can change according to AR.

This study confirms a positive statistical correlation of CFR with MR in addition to a positive relationship with AR.

An important cause of variances CFR across the world seems to be due to be previously underestimated unrecognized factor that is AR.

Recommendations: increased AR is a very high significantly associated possible predictor for increased MR and CFR. Measures focused on the reduction of AR will certainly reduce MR and CFR.

## Data Availability

I used publically available data.  Patients have been not involved

## Conflict of interest

There are no conflicts of interest worth to be mentioned.

Ethical approval was not required for this study, as we used publically available data, and patients were not involved.

## Acknowledgment

I am deeply grateful to Emeritus Professor Abdulkhaleq Abduljabbar Ali Ghalib Al-Naqeeb, Ph.D.in the Philosophy of Statistical Sciences - Medical - Health Technology college, Baghdad-Iraq, for his assistance in the data analysis, interpretation of the results and findings, and statistical revision of the paper.

---

**Table 1:** descriptive data regarding total CFR%, total attack rate (AR) / 10^4^, and total tests/million population inhabitants

| Country/territory Group | Total CFR% | Total MR/$10^4$ | Total (AR) / $10^4$ | Total tests/million |
| --- | --- | --- | --- | --- |
| **Group I (12 countries)** | **2.082** | **16.377** | **786.473** | **1,069,856.523** |
| **Group II (12 countries)** | **2.296** | **12.762** | **555.616** | **335,599.494** |
| **Group (III) (14 countries)** | **1.024** | **6.763** | **659.951** | **658,743** |

Country mortality rate groups: group I: $\geq 15$ death / $10^4$ population inhabitants; group II: $\geq 10$ -15 death / $10^4$ population inhabitants; and group III $<10$ death / $10^4$ population inhabitants. CFR; case fatality rate; AR: attack rate.

**Table 2:** Normal distribution function test (Goodness of fit test) for studied markers

| One-Sample Kolmogorov-Smirnov Test |  |  |  |  |
| --- | --- | --- | --- | --- |
| Markers | Statistics | Groups <sup>(**)</sup> |  |  |
|  |  | I | II | III |
| CFR % | No. | 12 | 12 | 16 |
|  | Kolmogorov-Smirnov Z | 0.589 | 0.481 | 0.509 |
|  | Asymptotic Sig. (2-tailed) | 0.878 | 0.975 | 0.958 |
|  | C.S. <sup>(*)</sup> | NS | NS | NS |
| AR X $10^4$ | No. | 12 | 12 | 16 |
|  | Kolmogorov-Smirnov Z | 0.591 | 0.848 | 0.739 |
|  | Asymptotic Sig. (2-tailed) | 0.876 | 0.468 | 0.646 |
|  | C.S. <sup>(*)</sup> | NS | NS | NS |
| Statistical Hypothesis: Ho: Markers are followed normal distribution function |  |  |  |  |
| Test distribution is Normal. |  |  |  |  |

<sup>(\*)</sup> NS: Non Sig. at $P>0.05$ .
<sup>(\*\*)</sup> Country mortality rate groups: group I: $\geq 15$ death / $10^4$ population inhabitants; group II: $\geq 10$ -15 death / $10^4$ population inhabitants; and group III $<10$ death / $10^4$ population inhabitants. CFR; case fatality rate; AR: attack rate.

**Table 3:** Summary Statistics concerning studied markers among different groups of countries regarding mortality rates

| Markers | Groups | No. | Mean | Std. D. | Std. E. | 95% C.I. for Mean |  | Min. | Max. |
| --- | --- | --- | --- | --- | --- | --- | --- | --- | --- |
|  |  |  |  |  |  | L.b. | U.b. |  |  |
| CFR % | I | 12 | 2.271 | 0.598 | 0.173 | 1.891 | 2.651 | 1.351 | 3.261 |
|  | II | 12 | 1.880 | 0.458 | 0.132 | 1.589 | 2.171 | 1.012 | 2.465 |
|  | III | 16 | 0.872 | 0.464 | 0.116 | 0.624 | 1.119 | 0.157 | 1.827 |
| AR X 10<sup>4</sup> | I | 12 | 881.7 | 294.9 | 85.1 | 694.3 | 1069.0 | 510.2 | 1286.6 |
|  | II | 12 | 734.1 | 242.5 | 70.0 | 580.0 | 888.2 | 517.5 | 1430.9 |
|  | III | 16 | 637.7 | 105.3 | 26.3 | 581.6 | 693.8 | 513.3 | 923.3 |

Country mortality rate groups: group I: $\geq 15$ death /10<sup>4</sup> population inhabitants; group II: $\geq 10$ -15 death / 10<sup>4</sup> population inhabitants; and group III <10 death /10<sup>4</sup> population inhabitants. CFR; case fatality rate; AR: attack rate.

**Table 4:** Testing equal variances and equal mean values for studied markers concerning different groups of countries classified according to different mortality rates /10^4^ population inhabitants

| Marker | Testing Homogeneity of Variances |  | ANOVA- Testing Equality of Means |  |
| --- | --- | --- | --- | --- |
|  | Levene Statistic | Sig. (*) | F-test | Sig. (*) |
| CFR% | 0.888 | 0.420 (NS) | 28.936 | 0.000 (HS) |
| AR/10<sup>4</sup> | 5.968 | 0.006 (HS) | 4.273 | 0.021 (S) |

(\*) HS: Highly Sig. at P<0.01; S: Sig. at P<0.05; NS: Non Sig. at P>0.05.

**Figure (1).**
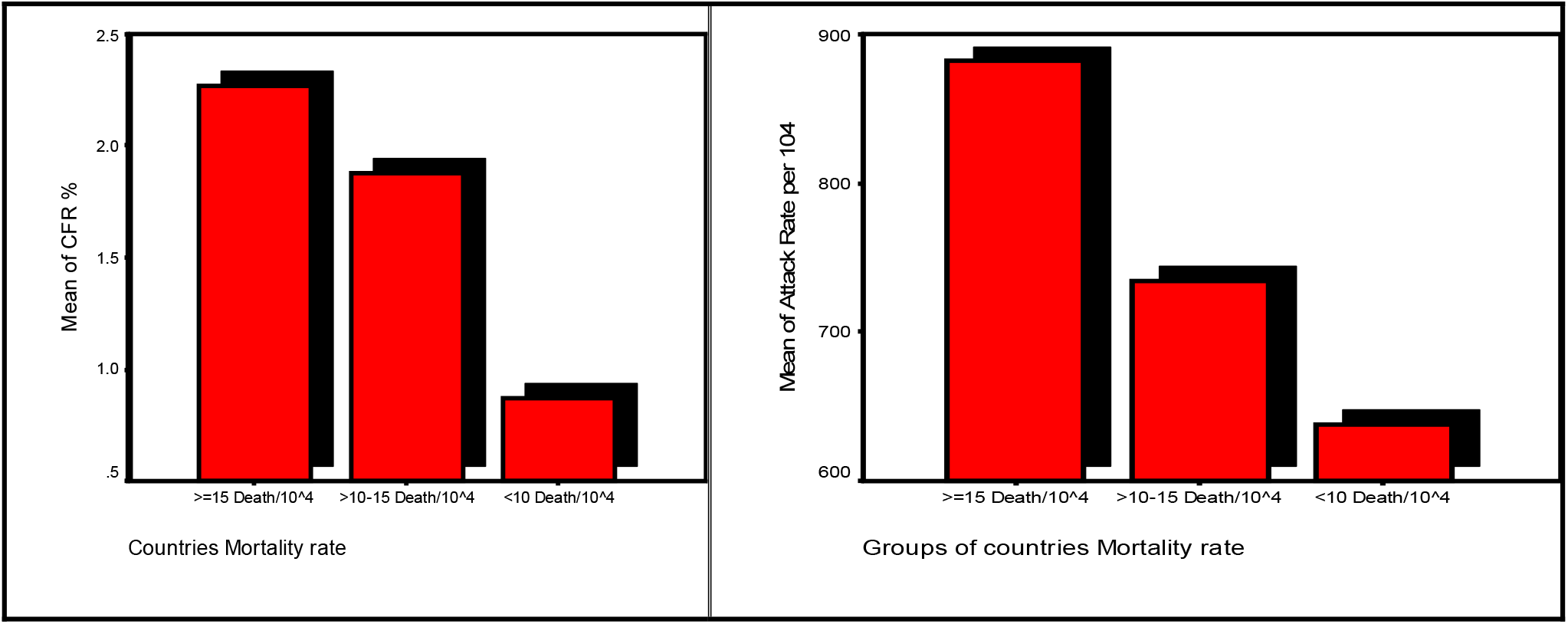
represent graphically plotting of bar chart regarding mean values studied marker’s readings distributed in different groups of countries regarding mortality rates. Country mortality rate groups: group I: ≥ 15 death /10^4^ population inhabitants; group II: ≥10-15 death / 10^4^ population inhabitants; and group III <10 death /10^4^ population inhabitants. CFR: case fatality rate; AR: attack rate

