## Supplementary file for "Covid-19 High Attack Rate Can Lead to High Case Fatality Rate"

**Appendices**

**Appendix 1: Basic collective data**

| **Country/ territory** | **cases** | **Deaths** | **CFR%** | **Population**  **As 3/10/2021)**  **2020 Population+**  **Growth rate**  **Up March10,2021** | **MR/10^4^** | **Attack rate (AR) / 104** | **Test/Million** | **No.tests** |
| --- | --- | --- | --- | --- | --- | --- | --- | --- |
| **Global** | **117,206,915** | **2,603,233** | **2.22** | **7,874,965,825** | **3.29768** | **148.834** | **-** |  |
| **Group of countries/ territories with mortality rate ≥ 15** | | | | | | | |  |
| **US** | **29,045,448** | **525,816** | **1.810** | **332,315,060** | **15.822** | **862.419** | **1,119,032** | **371,871,186** |
| **United Kingdom** | **4,235,989** | **125,032** | **2.952** | **68,105,326** | **18.359** | **621.976** | **1,440,364** | **98,096,460** |
| **Spain** | **3,164,982** | **71,727** | **2.266** | **46,749,627** | **15.3428** | **677.006** | **861,551** | **40,277,187** |
| **Italy** | **3,081,368** | **100,479** | **3.261** | **60,392,133** | **16.638** | **510.227** | **709,809** | **42,866,880** |
| **Czechia** | **1,335,815** | **22,147** | **1.658** | **10,720,144** | **20.659** | **1,246.079** | **843,453** | **9,041,938** |
| **Portugal** | **811,306** | **16,595** | **2.045** | **10,177,199** | **16.306** | **797.180** | **828,375** | **8,430,537** |
| **Slovenia** | **196,421** | **3,904** | **1.988** | **2,078,724** | **18.781** | **944.911** | **456,949** | **949,871** |
| **Macedonia** | **108,350** | **3,228** | **2.979** | **2,082,997** | **15.496** | **520.164** | **263,143** | **548,126** |
| [**Gibraltar**](https://www.worldometers.info/coronavirus/country/gibraltar/) | **4,255** | **93** | **2.186** | [**33,684**](https://www.worldometers.info/world-population/gibraltar-population/) | **27.609** | **1,263.211** | **5,775,383** | **194,538** |
| **San Marino** | **3,984** | **77** | **1.933** | **34,017** | **22.636** | **1,171.179** | **1,339,406** | **45,563** |
| **Montenegro** | **80,803** | **1,092** | **1.351** | **628,053** | **17.387** | **1,286.563** | **472,613** | **296,826** |
| **Belgium** | **789,008** | **22,292** | **2.825** | **11,618,642** | **19.186** | **679.088** | **893,719** | **10,383,801** |
| **Total** | **42,857,729** | **892,482** | **2.082** | **544,935,606** | **16.377** | 786.473 | **1,069,856.523** | **583,002,913** |
| **Mean 1** |  |  | **2.271** |  |  | **881.667** | **/ While 15,003,797/12=1,250,316.4** |  |
| **Group of countries/ territories with mortality rate ≥ 10-15** | | | | | | | |  |
| **Brazil** | **11,051,665** | **266,398** | **2.410** | **213,552,909** | **12.474** | **517.514** | **133,897** |  |
| **Andorra** | **11,069** | **112** | **1.012** | **77,355** | **14.479** | **1,430.935** | **2,502,844** |  |
| **France** | **3,992,755** | **89,455** | **2.240** | **65,376,867** | **13.682** | **610.729** | **850,494** |  |
| **Luxembourg** | **56,822** | **668** | **1.176** | **631,988** | **10.569** | **899.099** | **3,467,136** |  |
| **Liechtenstein** | **2,594** | **55** | **2.120** | **38,250** | **14.379** | **678.169** | **740,596** |  |
| **Armenia** | **175,198** | **3,232** | **1.845** | **2,966,657** | **10.894** | **590.556** | **255,358** |  |
| **Sweden** | **695,975** | **13,042** | **1.874** | **10,141,282** | **12.860** | **686.279** | **628,890** |  |
| **Switzerland** | **566,412** | **10,062** | **1.776** | **8,696,042** | **11.571** | **651.344** | **607,816** |  |
| **Panama** | **345,236** | **5,934** | **1.718** | **4,361,110** | **13.606** | **791.624** | **453,360** |  |
| **Slovakia** | **325,993** | **8,037** | **2.465** | **5,460,608** | **14.718** | **596.990** | **394,425** |  |
| **Croatia** | **247,099** | **5,621** | **2.275** | **4,089,002** | **13.747** | **604.301** | **341,537** |  |
| **Lithuania** | **202,900** | **3,341** | **1.647** | **2,699,927** | **12.374** | **751.501** | **812,342** |  |
| **Total** | **17,673,718** | **405,957** | **2.296** | **318,091,997** | **12.762** | **555.616** | **35,174.399** | **11,188,695** |
| **Mean** |  |  | **1.80** |  |  | **734.087** |  |  |
| **Group of countries / territories with mortality rate <10** | | | | | | | |  |
| **Netherlands** | **1,143,395** | **16,046** | **1.403** | **17,161,450** | **9.350** | **666.258** | **406,178** |  |
| **Qatar** | **167,888** | **264** | **0.157** | **2,913,792** | **0.906** | **576.183** | **566,655** |  |
| **Bahrain** | **127,255** | **473** | **0.372** | **1,734,048** | **2.728** | **733.860** | **1,852,557** |  |
| **Israel** | **807,755** | **5,926** | **0.734** | **8,748,382** | **6.774** | **923.319** | **1,378,194** |  |
| **Georgia** | **273,137** | **3,601** | **1.318** | **3,982,818** | **9.041** | **685.788** | **723,652** |  |
| **Estonia** | **77,491** | **669** | **0.863** | **1,325,637** | **5.046** | **584.556** | **755,538** |  |
| [**French Polynesia**](https://www.worldometers.info/coronavirus/country/french-polynesia/) | **18,495** | **141** | **0.762** | [**282,024**](https://www.worldometers.info/world-population/french-polynesia-population/) | **5.000** | **655.795** | **93,449** |  |
| [**Mayotte**](https://www.worldometers.info/coronavirus/country/mayotte/) | **18,413** | **129** | **0.701** | [**277,296**](https://www.worldometers.info/world-population/mayotte-population/) | **4.652** | **664.019** | **517,746** |  |
| [**Aruba**](https://www.worldometers.info/coronavirus/country/aruba/) | **8,177** | **77** | **0.9417** | [**107,078**](https://www.worldometers.info/world-population/aruba-population/) | **7.191** | **763.648** | **1,030,856** |  |
| **Serbia** | **494,106** | **4,599** | **0.930** | **8,709,651** | **5.280** | **567.309** | **352,584** |  |
| **Austria** | **479,391** | **8,757** | **1.827** | **9,031,309** | **9.696** | **530.810** | **639,106** |  |
| **Lebanon** | **397,887** | **5,134** | **1.290** | **6,777,741** | **7.575** | **587.049** | **467,432** |  |
| **Monaco** | **2,028** | **26** | **1.282** | **39,511** | **6.580** | **513.275** | **488,382** |  |
| [**French Guiana**](https://www.worldometers.info/coronavirus/country/french-guiana/) | **16,693** | **87** | **0.5211** | [**303,947**](https://www.worldometers.info/world-population/french-guiana-population/) | **2.862** | **549.208** | **504,914** |  |
| [**Turks and Caicos**](https://www.worldometers.info/coronavirus/country/turks-and-caicos-islands/) | **2,177** | **15** | **0.689** | [**39,076**](https://www.worldometers.info/world-population/turks-and-caicos-islands-population/) | **3.839** | **557.119** | **444,416** |  |
| [**St. Barth**](https://www.worldometers.info/coronavirus/country/saint-barthelemy/) | **638** | **1** | **0.157** | [**9,898**](https://www.worldometers.info/world-population/saint-barthelemy-population/) | **1.010** | **644.574** | **1,931,097** |  |
| Total | **4,034,926** | **41,346** | **1.024** | 61,139,711 | **6.763** | **659.951** | **198,770.255** | **12,152,756** |
| Mean |  |  | **0.8717** |  |  | **637.673125** |  |  |

**Appendix 2: total group statistics ( derived from appendix 1)**

| **Country/ territory** | **cases** | **Deaths** | **CFR%** | **Population**  **As 3/10/2021)**  **2020 Population+**  **Growth rate**  **Up March10,2021** | **MR/10^4^** | **Attack rate (AR) / 10^4^** | **Test/Million** | **No.tests** |
| --- | --- | --- | --- | --- | --- | --- | --- | --- |
| **Global** | **117,206,915** | **2,603,233** | **2.22** | **7,874,965,825** | **3.29768** | **148.834** | **-** |  |
| **Group of countries/ territories with mortality rate ≥ 15** | | | | | | | |  |
| **US** | **29,045,448** | **525,816** | **1.810** | **332,315,060** | **15.822** | **862.419** | **1,119,032** | **371,871,186** |
| **United Kingdom** | **4,235,989** | **125,032** | **2.952** | **68,105,326** | **18.359** | **621.976** | **1,440,364** | **98,096,460** |
| **Spain** | **3,164,982** | **71,727** | **2.266** | **46,749,627** | **15.3428** | **677.006** | **861,551** | **40,277,187** |

**Appendix 3: CFR and AR for group of countries / territories with mortality rate ≥ 15 death / 10^4^ population inhabitants**

| **Country** | **CFR%** | **Attack Rate (AR) per 10^4^** |
| --- | --- | --- |
| **US** | **1.810** | **862.419** |
| **United Kingdom** | **2.952** | **621.976** |
| **Spain** | **2.266** | **677.006** |
| **Italy** | **3.261** | **510.227** |
| **Czechia** | **1.658** | **1,246.079** |
| **Portugal** | **2.045** | **797.180** |
| **Slovenia** | **1.988** | **944.911** |
| **Macedonia** | **2.979** | **520.164** |
| [**Gibraltar**](https://www.worldometers.info/coronavirus/country/gibraltar/) | **2.186** | **1,263.211** |
| **San Marino** | **1.933** | **1,171.179** |
| **Montenegro** | **1.351** | **1,286.563** |
| **Belgium** | **2.825** | **679.088** |
| **Mean** | **2.271** | **881.667** |

**Appendix 4: CFR and AR for group of countries/ territories with mortality rate ≥ 10-15 death / 10^4^ population inhabitants**

| **Country / territory** | **CFR%** | **Attack Rate (AR) per 10^4^** |
| --- | --- | --- |
| **Brazil** | **2.410** | **517.514** |
| **Andorra** | **1.012** | **1,430.935** |
| **France** | **2.240** | **610.729** |
| **Luxembourg** | **1.176** | **899.099** |
| **Liechtenstein** | **2.120** | **678.169** |
| **Armenia** | **1.845** | **590.556** |
| **Sweden** | **1.874** | **686.279** |
| **Switzerland** | **1.776** | **651.344** |
| **Panama** | **1.718** | **791.624** |
| **Slovakia** | **2.465** | **596.990** |
| **Croatia** | **2.275** | **604.301** |
| **Lithuania** | **1.647** | **751.501** |
| **Mean** | **1.80** | **734.087** |

**Appendix 5: CFR and AR for group of countries/ territories with mortality rate <10 death / 10^4^**

| **Country / territory** | **CFR%** | **Attack Rate (AR)per 10^4^** |
| --- | --- | --- |
| **Netherlands** | **1.403** | **666.258** |
| **Qatar** | **0.157** | **576.183** |
| **Bahrain** | **0.372** | **733.860** |
| **Israel** | **0.734** | **923.319** |
| **Georgia** | **1.318** | **685.788** |
| **Estonia** | **0.863** | **584.556** |
| [**French Polynesia**](https://www.worldometers.info/coronavirus/country/french-polynesia/) | **0.762** | **655.795** |
| [**Mayotte**](https://www.worldometers.info/coronavirus/country/mayotte/) | **0.701** | **664.019** |
| [**Aruba**](https://www.worldometers.info/coronavirus/country/aruba/) | **0.9417** | **763.648** |
| **Serbia** | **0.930** | **567.309** |
| **Austria** | **1.827** | **530.810** |
| **Lebanon** | **1.290** | **587.049** |
| **Monaco** | **1.282** | **513.275** |
| [**French Guiana**](https://www.worldometers.info/coronavirus/country/french-guiana/) | **0.5211** | **549.208** |
| [**Turks and Caicos**](https://www.worldometers.info/coronavirus/country/turks-and-caicos-islands/) | **0.689** | **557.119** |
| [**St. Barth**](https://www.worldometers.info/coronavirus/country/saint-barthelemy/) | **0.157** | **644.574** |
| Mean | **0.8717** | **10,202.77/16= 637.673125** |

**Appendix 6**

**A-References for population data: general**

1- **UN. Department of Economic and Social Affairs. Population. *2019 Revision* of *World Population Prospects* . 2020 estimates in attached Excel file. Accessed March 3,2021**
2- **World bank :https://data.worldbank.org/indicator/SP.POP.TOTL**
3- [**2021 World Population by Country (worldpopulationreview.com)**](https://worldpopulationreview.com/)

**B-Country specific**

**Kosovo : United Nations Department of Economic and Social Affairs: Population Division.**

1. **Gaza;** [**Palestinian Central Bureau of Statistics**](http://www.pcbs.gov.ps/Portals/_Rainbow/Documents/gza.htm) **- Localities in Gaza Governorate by type of locality and population estimates - 2007-2016**
2. [**World Urbanization Prospects**](https://esa.un.org/unpd/wup/) **- United Nations population estimates and projections of major Urban Agglomerations**

**Appendix 7:COVID-19 data**

1- [**COVID-19 Map - Johns Hopkins Coronavirus Resource Center (jhu.edu)**](https://coronavirus.jhu.edu/map.html)
2-
3-
4. [4- ](   WHO COVID-19 Explorer. Geneva: World Health Organization, 2020. Available online: https://worldhealthorg.shinyapps.io/covid/ (last cited: [date]).)**[WHO COVID-19 Explorer. Geneva: World Health Organization, 2020. Available online: https://worldhealthorg.shinyapps.io/covid/ (last cited: [date]).](   WHO COVID-19 Explorer. Geneva: World Health Organization, 2020. Available online: https://worldhealthorg.shinyapps.io/covid/ (last cited: [date]).)**
